# Dengue Hospitalizations in Brazil: Forecasting with Climatic and Physicians’ Digital Search Data under Real-World Reporting Delays

**DOI:** 10.64898/2026.01.12.26343977

**Authors:** Dayanna Quintanilha Palmer, Marcela Motta, Eduardo Cardoso de Moura, Danielly Xavier, Guilherme Schittine, Angélica Caseri, Ronaldo Altenburg Gismondi

**Author notes:** Corresponding author (DQP).

## Abstract

Timely forecasting of dengue hospitalizations is essential for public health preparedness but is frequently limited by delays in official reporting systems. Climatic conditions strongly influence dengue transmission, yet hospitalization data often become available weeks after patient admission, reducing their value for early response. Digital information generated during clinical practice may provide a more timely signal of emerging disease activity.

This study evaluates whether integrating climate data with real-time records of physicians’ searches for dengue-related information improves short-term forecasts of dengue hospitalizations in Brazil under both ideal and realistic reporting conditions. Weekly hospitalization counts, weather indicators, and anonymized physician search data from a widely used clinical decision-support platform were combined to generate forecasts across multiple geographic regions. Model performance was compared under two scenarios: one assuming immediate availability of hospitalization data and another incorporating typical reporting delays.

When hospitalization data were timely, climate-based models achieved the highest predictive accuracy. Under realistic reporting delays, however, models incorporating physicians’ search behavior consistently outperformed approaches relying solely on climate information or hospitalization history. In several regions, increases in physician search activity preceded or coincided with rises in hospital admissions, indicating early clinical engagement with dengue cases.

These findings indicate that physician search behavior constitutes a valuable real-time indicator of dengue activity. Integrating digital clinical behavior with climate data enhances forecasting performance under real-world reporting constraints and may strengthen early-warning systems and public health decision-making for dengue and other climate-sensitive diseases.

**Author Summary:** Dengue is a major public health challenge in Brazil, where large outbreaks place sudden pressure on health services. Although climate conditions influence dengue transmission, public health responses often rely on hospitalization data that become available only weeks or months after patients are admitted, limiting the ability to act early. In this study, we explored whether real-time digital information generated by physicians could help overcome this delay.

We combined weather data with anonymized records of physicians’ searches for dengue-related information within a widely used clinical decision-support platform in Brazil. We then tested whether these digital signals could improve short-term forecasts of dengue hospitalizations across different regions of the country, especially when official hospital data were delayed.

We found that climate patterns were strong predictors when hospitalization data were timely. However, under realistic reporting delays, models that incorporated physicians’ search behavior produced more accurate forecasts. These findings show that digital clinical behavior can provide early insight into rising disease activity and support more timely public health responses.

## 1. Introduction

Dengue virus (DENV) is an arbovirus with four distinct serotypes (DENV-1 to DENV-4), primarily transmitted by *Aedes* mosquitoes, especially *Aedes aegypti*. While primary infection may be asymptomatic or present with mild symptoms, severe cases can progress to dengue hemorrhagic fever and dengue shock syndrome, characterized by coagulopathy and increased vascular permeability, which can ultimately be fatal if not promptly recognized and managed (1,2). In 2024, Brazil recorded approximately 6.42 million probable dengue cases, representing a 327% increase compared to 2023, when 1.51 million probable cases were reported (3)

Several factors may contribute to the rise in dengue cases, with climate change emerging as a key driver of mosquito-borne disease dynamics. Temperatures exceeding regional threshold levels accelerate the life cycle of dengue vectors (*Aedes aegypti*, and *Aedes albopictus*), increase dengue virus proliferation and mosquito bite frequency, and shorten the extrinsic incubation period (4,5). Higher humidity and precipitation have been linked to increased dengue transmission, as they create favorable conditions for virus replication and spread (4,6). Furthermore, socio-economic factors play an important role in shaping dengue transmission patterns, contributing to a complex system that influences the probability of future outbreaks (7,8).

Accurately mapping geographic distribution and predicting outbreaks is essential to guide resource allocation and enable timely public health interventions (6,9). Several predictive models have been proposed, but most focus exclusively on climate variables and often neglect the effect of time lags in transmission dynamics (8). In addition to these analytical lags, forecasting efforts are also constrained by reporting delays in official hospitalization and case-notification systems, which reduce the timeliness and operational usefulness of surveillance data (10). Moreover, digital behavioral data from physicians—such as clinical search activity within medical decision-support platforms—have not yet been explored in the context of dengue modeling in Brazil, despite their potential to provide early indicators of clinical demand and contribute to real time decision making.

Integrating meteorological and non-climatic factors may substantially improve predictive accuracy (11,12). In this context, digital health data provides a novel opportunity for syndromic surveillance (13,14). Patterns of access to medical applications can reflect real-time shifts in medical concerns and population health trends. In Brazil, the Afya Whitebook® application, a widely used digital health tool among physicians, captures search behaviors, including queries related to dengue. In a previous study, these digital traces showed significant correlation with hospitalizations for dengue, supporting their use as a proxy of clinical demand (15).

Building upon this rationale, the present study proposes a predictive framework to forecast weekly dengue hospitalizations in Brazil by integrating climatic variables, modeled with lagged predictors to capture delayed environmental effects, and digital health indicators derived from physicians’ search behavior within the Afya Whitebook® platform. To address hospitalization reporting delays, we developed two modeling scenarios: an ideal-data model, assuming real-time data availability, and a real-world model, which incorporates the typical delay observed in hospitalization record updates. By comparing these model configurations, we aim to evaluate how incorporating realistic data latency and temporal dependencies can enhance the accuracy and operational relevance of short-term dengue forecasting.

## 2. Methods

### 2.1 Study design

We developed a forecasting framework based on Long Short-Term Memory (LSTM) neural networks to predict dengue-related hospitalizations in Brazil at the Immediate Geographic Region (IGR) level. **Figure 1** provides an overview of the analytical workflow, which includes:

(i) integration of climate, physicians’ clinical search, and hospitalization datasets from public service;
(ii) preprocessing steps such as weekly aggregation, imputation of missing values, and consolidation at the IGR level;
(iii) derivation of additional climate indicators;
(iv) variable screening using LSTM-based relevance evaluation; and
(v) model training and interpretability using SHAP (SHapley Additive Explanations) values.

**Figure 1.**
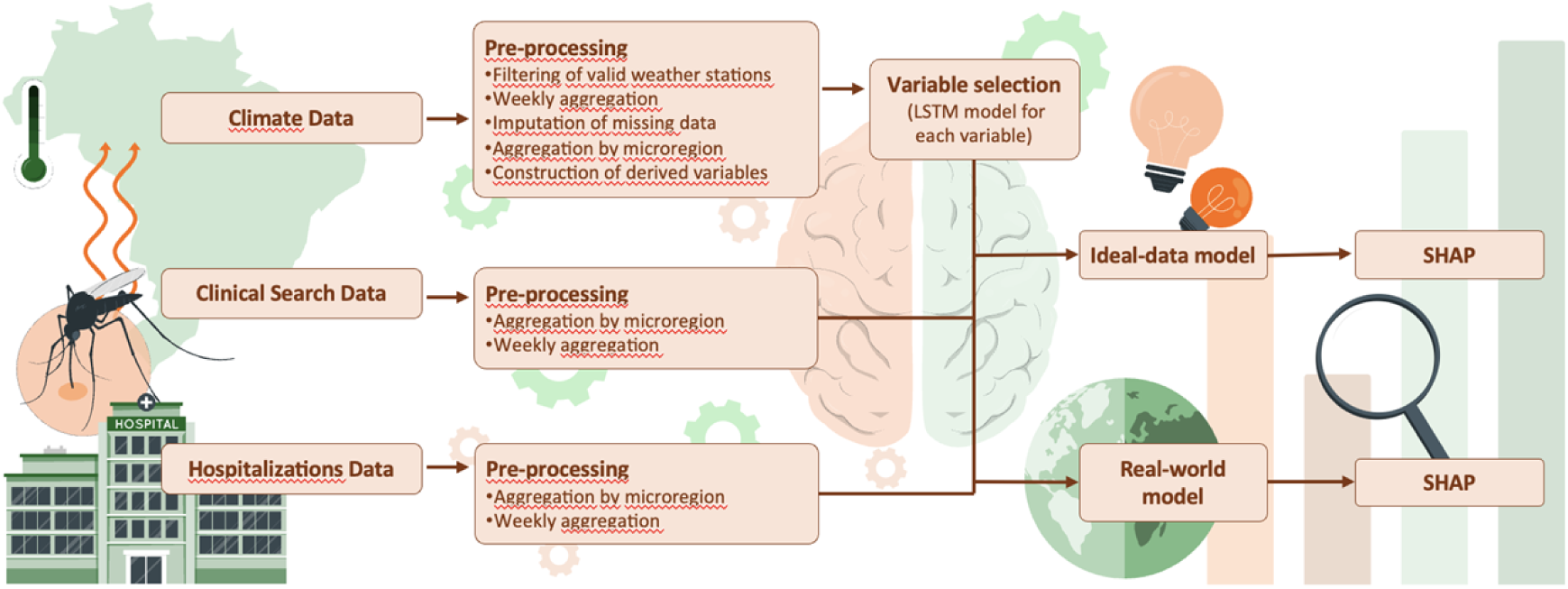
Workflow of the analytical pipeline, including preprocessing of weather station data, weekly aggregation, imputation of missing values, microregion-level aggregation, derivation of climate variables, LSTM-based variable selection, and SHAP analyses for both ideal-data and real-world scenarios.

Given the temporal nature of the data and the need to capture both short and long-term dependencies in dengue transmission, we employed LSTM networks, a type of recurrent neural network specifically designed to handle sequential data with temporal correlations(16). LSTM models have been successfully applied in infectious disease forecasting, including dengue(11), demonstrating strong performance in predicting case trends based on climatic and other exogenous variables.

The modeling strategy was designed to generate 8-week-ahead forecasts, a horizon chosen to reflect both epidemiological relevance and operational usefulness for public health planning. To capture the intrinsic temporal structure of dengue transmission, we implemented a rolling sliding-window approach, in which recent historical sequences were used to predict subsequent hospitalization counts. This setup enabled the model to learn meaningful temporal patterns and produce actionable short- to medium-term forecasts.

### 2.2 Study site

According to the Brazilian Institute of Geography and Statistics (IBGE), an IGR corresponds to a group of neighboring municipalities organized around one or more urban centers that function as local hubs for goods, services, and labor. These regions reflect short-distance functional relationships, particularly commuting flows and the daily circulation of people and economic activities.

The current territorial division, comprises 510 IGRs, that represent an intermediate geographic scale between municipalities and states, ensuring spatial coherence for socioeconomic, environmental, and health analyses (17).

### 2.3 Data sources

#### 2.3.1 Hospitalizations Data

The SIH/SUS database, maintained by the Department of Informatics of the Unified Health System (DATASUS), provides anonymized, publicly available information on hospital admissions throughout Brazil (18). Data are updated monthly, with a typical release delay of one to two months after hospital care. In accordance with Portaria SAES n° 1.110/2021 of the Brazilian Ministry of Health, the final consolidation of hospitalization data within the SIH/SUS may occur up to four months after the reference month, due to the administrative cycles of data submission, verification, and correction of hospitalization authorizations (10).

#### 2.3.2 Clinical Search Data

Afya Whitebook® is a widely used clinical decision-support platform among healthcare professionals in Brazil. The platform provides structured, evidence-based content such as disease overviews, diagnostic algorithms, therapeutic guidelines, drug dosing information, and clinical calculators. Optimized for mobile and desktop use, it supports real-time clinical decision-making across a variety of care settings. Although its primary audience is licensed physicians, the user base also includes residents, medical students, and other allied health professionals. For this study, only search data generated by verified licensed physicians with active professional credentials were included. In Brazil, there were 597,428 practicing physicians as of January 2024, according to the recent edition of Demografia Médica(19). In comparison, the Afya Whitebook® platform registered approximately 150,000 monthly active physician users by the end of the same year (20).

The Afya Whitebook® research database contains metadata on user search behavior, collected in real time and accessible retrospectively for research purposes via a structured query interface. All data are anonymized to ensure confidentiality and compliance with ethical standards, preventing the identification of individual healthcare professionals.

#### 2.3.3 Climate data

Daily observations of precipitation, maximum, mean, and minimum temperature, and mean relative humidity were obtained from the Brazilian National Institute of Meteorology (Instituto Nacional de Meteorologia – INMET) automatic weather stations distributed throughout Brazil(21).

### 2.4 Data pre-processing

Climate data underwent several cleaning and harmonization steps. Dates were converted to datetime format, variable names standardized, and station metadata (name, geographic coordinates, operational period) appended to each record. Only observations between 2021 and 2024 were retained.

Stations with more than 30 consecutive days of missing data were excluded, following the revised criterion established after model calibration. Among IGRs containing multiple meteorological stations, only the station with the highest percentage of completeness was retained.

Daily measures were aggregated into weekly series, with averages computed for temperature and humidity, and weekly sums for precipitation. Missing weekly values were imputed using a centered moving average, calculated with two weeks forward and two weeks backward (minimum of one available neighbor), ensuring temporal continuity of the time series.

### 2.5 Study variables

The primary outcome of interest was the weekly number of hospitalizations for dengue fever extracted from the SIH/SUS database. Predictor variables encompassed both meteorological and digital health domains. Meteorological predictors included lagged indicators of precipitation, temperature, and humidity (1–4 weeks), alongside a structured derivation process that resulted in a final set of 42 climate-related variables, including weekly change flags, interaction terms, extreme climate indicators, seasonal dummies, temperature categories, and precipitation occurrence and patterns. In addition, clinical search data were incorporated through weekly counts of physician searches for dengue in Afya Whitebook®, restricted to verified licensed physicians. Together, these variables were selected to capture the combined influence of climatic fluctuations and real-time medical information-seeking behavior on dengue-related hospitalizations. All derived variables are summarized in Table 1, and their detailed definitions and computation procedures are provided in the Supplementary Material (Table S1).

**Table 1.**
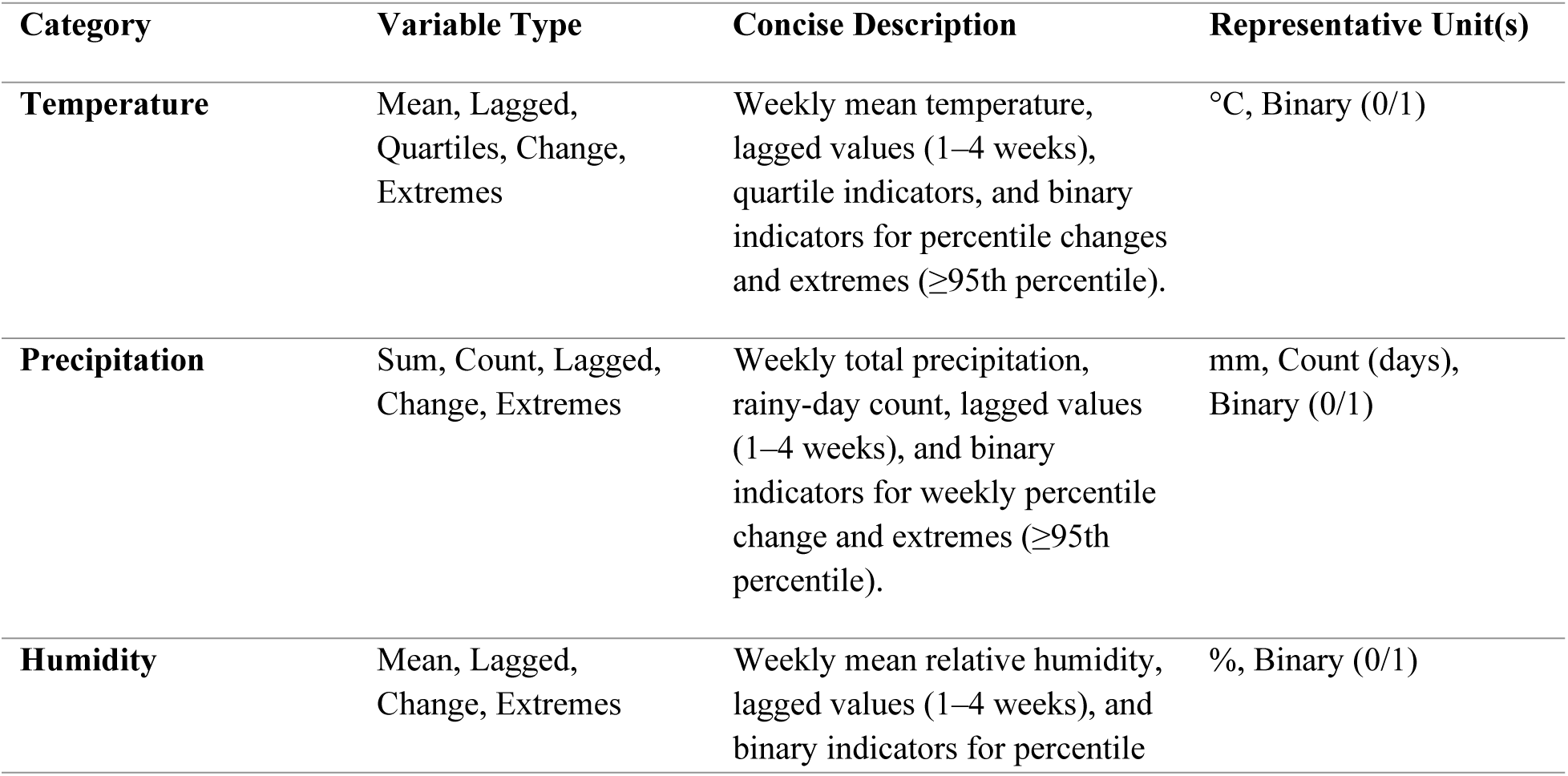
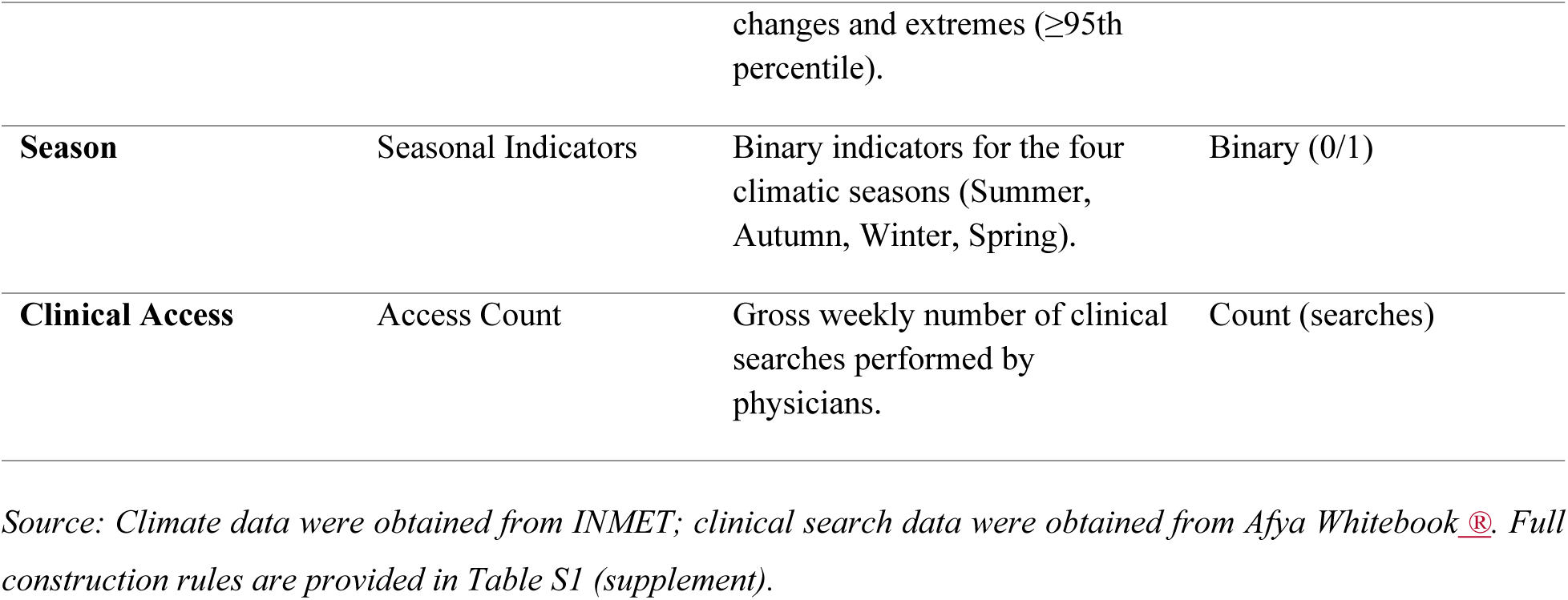
Summary of the categories and types of variables included in the models.

| Category | Variable Type | Concise Description | Representative Unit(s) |
| --- | --- | --- | --- |
| **Temperature** | Mean, Lagged, Quartiles, Change, Extremes | Weekly mean temperature, lagged values (1–4 weeks), quartile indicators, and binary indicators for percentile changes and extremes ($\geq 95$th percentile). | °C, Binary (0/1) |
| **Precipitation** | Sum, Count, Lagged, Change, Extremes | Weekly total precipitation, rainy-day count, lagged values (1–4 weeks), and binary indicators for weekly percentile change and extremes ($\geq 95$th percentile). | mm, Count (days), Binary (0/1) |
| **Humidity** | Mean, Lagged, Change, Extremes | Weekly mean relative humidity, lagged values (1–4 weeks), and binary indicators for percentile changes and extremes ( $\geq 95$ th percentile). | %, Binary (0/1) |
| **Season** | Seasonal Indicators | Binary indicators for the four climatic seasons (Summer, Autumn, Winter, Spring). | Binary (0/1) |
| **Clinical Access** | Access Count | Gross weekly number of clinical searches performed by physicians. | Count (searches) |

*Source: Climate data were obtained from INMET; clinical search data were obtained from Afya Whitebook <sup>®</sup>. Full construction rules are provided in Table S1 (supplement).*

### 2.6 Feature selection

Given the presence of 42 climatic input variables derivations, we sought to reduce model dimensionality and avoid multicollinearity by selecting only one variable from each climatic category—humidity, temperature, precipitation, and seasonality. To achieve this, we employed a variable-by-variable evaluation approach, in which the model was trained using the hospitalization variable alongside a single climatic predictor at a time. For each category, the climatic variable yielding the lowest RMSE was retained. As a result, each IGR retained four climatic input variables, one from each category.

Feature importance was further assessed using SHAP values, which enabled interpretation of the most influential predictors per IGR and quantification of the relative contribution of climatic versus digital health variables.

### 2.7 Data analysis

Hospitalization, digital query, and climatic datasets were merged at the IGR–week level to form a unified analytic panel. Exploratory analyses included visualization of temporal trends and descriptive statistics across regions. Forecasting was performed using a LSTM neural network trained on weekly climatic and digital health predictors. To account for temporal dependence, the dataset was split chronologically into training and testing sets, with the training period spanning 2021–2023 and the testing period covering 2024.

All data preprocessing and modeling were conducted in a Python-based analytical environment (22). Hyperparameters were optimized using a grid search evaluating different architectural configurations, including the number of hidden layers (2–4) and hidden units (20 or 40). The final model employed an input window of 24 weeks (seq_len = 24), a prediction horizon of 8 weeks (output_size = 8), a learning rate of 0.01, and a MSE loss function. Each configuration was trained in triplicate to account for stochastic variability during optimization. Model performance was assessed using the RMSE, the MSE, the Mean Absolute Error (MAE), and the coefficient of determination (R²).

These metrics were computed as follows:

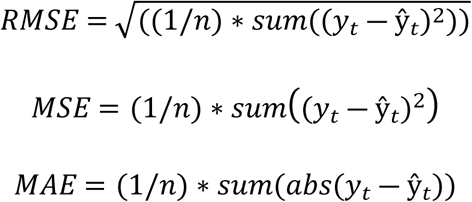

To compute the coefficient of determination, we used the Residual Sum of Squares (RSS) and the Total Sum of Squares (TSS):

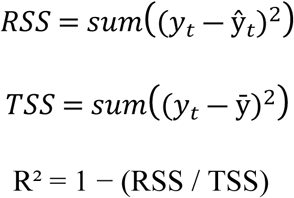

These quantities quantify both the magnitude of prediction errors and the proportion of variance in weekly hospitalizations explained by the forecasting model.

To evaluate the relative contribution of different data domains, five model structures were compared:

- **Hospitalization-only model (LSTM – Hospitalization):** Trained exclusively on past hospitalization counts to capture the intrinsic temporal dynamics of dengue without external predictors.
- **Clinical-search model (LSTM – Hospitalization + Clinical Search):** Trained using hospitalization time series combined with Afya Whitebook® physician search data.
- **Climate model (LSTM – Hospitalization + Climate):** Trained using hospitalization data and climatic predictors, selecting the best-performing variable from each domain (temperature, humidity, precipitation).
- **Integrated model (LSTM – Hospitalization + Clinical Search + Climate):** Combines hospitalization data, climatic indicators, and real-time physician search behavior, restricted to the most informative variable per domain.
- **SARIMAX model (SARIMAX – Hospitalization):** A classical benchmark trained only on hospitalization counts, used as a baseline for comparative performance assessment (23).

In addition, two complementary versions were implemented:

- **Ideal-data model:** predictors and hospitalization data were temporally aligned.
- **Real-world model:** hospitalization data were shifted by up to eight weeks (approximately two months).

The two-month delay window was empirically defined based on the average reporting lag observed in the hospitalization dataset, ensuring that the model reflected the actual latency pattern present in official records. Performance differences between models were statistically compared using paired t-tests across triplicate RMSE results.

A walk-forward expanding-window validation approach was adopted during training to mimic real-time forecasting and assess model robustness across temporal segments. Predicted versus observed hospitalization counts were visually inspected to evaluate both numerical accuracy and temporal alignment.

#### 2.7.1 LSTM model Architeture

LSTM networks are a special type of recurrent neural network (RNN) designed to overcome the vanishing and exploding gradient problem in long sequences (16). An LSTM unit contains a memory cell with a constant error carousel (CEC) that preserves information across time steps, while multiplicative gates regulate information flow. Gers, Schmidhuber, and Cummins (2000) extended the model with the forget gate, allowing adaptive resetting of memory contents ((24).

In our study, the LSTM architecture was adapted to model the temporal dynamics of dengue hospitalizations in Brazil. The input, forget, and output gates controlled the flow of information across time, while the memory cell preserved long-range dependencies, allowing the model to integrate both climatic fluctuations and digital health signals. Following this principle, we structured the implementation to use weekly lagged climate indicators, extreme event markers, and physician search behavior from Afya Whitebook® as inputs. Through a sliding-window training strategy, the LSTM was able to capture short-term variations and longer seasonal patterns, providing forecasts at the microregion level that aligned with the theoretical design of the architecture.

### 2.8 Ethics approval

The research complies with the ethical principles established by Resolution CNS 466/12 of the Brazilian National Health Council. All data used were anonymized and derived from public or internal databases, with no individual-level identifiers or direct participant interaction.

The research protocol was reviewed and approved by the Research Ethics Committee of the Centro Universitário Presidente Tancredo de Almeida Neves (UNIPTAN), under CAAE 81123624.4.0000.9667 and Opinion number 6.944.607. The Committee granted a waiver of informed consent given the exclusive use of anonymized secondary data.

## 3. Results

### 3.1 Characterization of included Immediate Geographic Regions and data coverage

The initial dataset encompassed 510 IGRs with dengue hospitalizations recorded in the SIH/SUS system between 2021 and 2024. After integrating hospitalization, climate, and Afya Whitebook® digital search data, sequential filtering steps were applied to ensure data completeness and model stability.

First, INMET weather stations with more than 30 consecutive missing days were excluded, and duplicated stations within the same IGR were resolved by retaining those with the highest proportion of complete observations. Clinical search data were available for 46 IGRs; however, only those with consistent weekly dengue-related searches and valid hospitalization data were retained. Additional exclusions occurred during preliminary modeling, in which IGRs whose base LSTM models produced non-positive R² values, reflecting insufficient temporal structure for forecasting, were removed.

After all filtering stages, a final set of 27 epidemiologically meaningful IGRs remained: Alegre, Belo Horizonte, Campina Grande, Campos Dos Goytacazes, Catalão, Cruz Alta, Distrito Federal, Frederico Westphalen, Ijui, Juiz De Fora, Linhares, Marília, Maringá, Oliveira, Passo Fundo, Passos, Pirapora, Porto Alegre, Ribeirão Preto, Rio De Janeiro, Salvador, Santa Cruz Do Sul, Santa Maria, São Miguel Do Oeste, São Paulo, Uberaba, Uberlândia. These regions displayed sufficient data density across all domains, enabling robust multi-step forecasting.

### 3.2 Spatiotemporal Trends in Hospitalization and Clinical Search Data (2021–2024)

Hospitalizations increased across most IGRs between 2021 and 2024 (Table 2). Smaller-population regions such as Frederico Westphalen, Oliveira, São Miguel do Oeste, and Uberlândia exhibited high hospitalization rates, frequently surpassing 200 to 600 per 100,000 inhabitants in peak year, while large metropolitan centers, including São Paulo, and Rio de Janeiro, reported low hospitalization rates, even though they presented the highest absolute numbers.

**Table 2.** Hospitalizations and Rates per 100000 inhabitants in 27 Selected Brazilian Immediate Geographic Regions, 2021–2024.

| Immediate Geographic Regions | Population | 2021 |  | 2022 |  | 2023 |  | 2024 |  |
| --- | --- | --- | --- | --- | --- | --- | --- | --- | --- |
|  |  | N | Rate | N | Rate | N | Rate | N | Rate |
| **Alegre** | 193922 | 4 | 2.06 | 19 | 9.80 | 166 | 85.60 | 245 | 126.34 |
| **Belo Horizonte** | 5045254 | 46 | 0.91 | 117 | 2.32 | 996 | 19.74 | 8119 | 160.92 |
| **Campina Grande** | 938502 | 129 | 13.75 | 275 | 29.30 | 50 | 5.33 | 319 | 33.99 |
| **Campos Dos Goytacazes** | 631164 | 4 | 0.63 | 27 | 4.28 | 321 | 50.86 | 726 | 115.03 |
| **Catalão** | 170345 | 45 | 26.42 | 215 | 126.21 | 36 | 21.13 | 279 | 163.79 |
| **Cruz Alta** | 138860 | 1 | 0.72 | 4 | 2.88 | 81 | 58.33 | 112 | 80.66 |
| **Distrito Federal** | 2817381 | 411 | 14.59 | 1292 | 45.86 | 1005 | 35.67 | 7408 | 262.94 |
| **Frederico Westphalen** | 119099 | 32 | 26.87 | 228 | 191.44 | 18 | 15.11 | 742 | 623.01 |
| **Ijuí** | 203825 | 2 | 0.98 | 27 | 13.25 | 55 | 26.98 | 373 | 183.00 |
| **Juiz De Fora** | 711921 | 9 | 1.26 | 17 | 2.39 | 122 | 17.14 | 1052 | 147.77 |
| **Linhares** | 333129 | 62 | 18.61 | 27 | 8.10 | 174 | 52.23 | 288 | 86.45 |
| **Marília** | 380834 | 78 | 20.48 | 182 | 47.79 | 110 | 28.88 | 245 | 64.33 |
| **Maringá** | 811915 | 43 | 5.30 | 477 | 58.75 | 278 | 34.24 | 950 | 117.01 |
| **Oliveira** | 115926 | 1 | 0.86 | 62 | 53.48 | 93 | 80.22 | 469 | 404.57 |
| **Passo Fundo** | 278694 | 3 | 1.08 | 7 | 2.51 | 49 | 17.58 | 108 | 38.75 |
| **Passos** | 258889 | 3 | 1.16 | 97 | 37.47 | 349 | 134.81 | 671 | 259.18 |
| **Pirapora** | 133939 | 1 | 0.75 | 11 | 8.21 | 146 | 109.00 | 296 | 221.00 |
| **Porto Alegre** | 2997642 | 6 | 0.20 | 595 | 19.85 | 310 | 10.34 | 1619 | 54.01 |
| **Ribeirão Preto** | 1446059 | 19 | 1.31 | 191 | 13.21 | 260 | 17.98 | 1101 | 76.14 |
| **Rio De Janeiro** | 11760550 | 21 | 0.18 | 151 | 1.28 | 1061 | 9.02 | 3095 | 26.32 |
| **Salvador** | 3485014 | 11 | 0.32 | 167 | 4.79 | 749 | 21.49 | 369 | 10.59 |
| **Santa Cruz Do Sul** | 364672 | 80 | 21.94 | 27 | 7.40 | 12 | 3.29 | 149 | 40.86 |
| **Santa Maria** | 461725 | 1 | 0.22 | 4 | 0.87 | 130 | 28.16 | 157 | 34.00 |
| **São Miguel Do Oeste** | 172124 | 11 | 6.39 | 533 | 309.66 | 34 | 19.75 | 406 | 235.88 |
| **São Paulo** | 20731920 | 125 | 0.60 | 141 | 0.68 | 207 | 1.00 | 7436 | 35.87 |
| **Uberaba** | 454392 | 19 | 4.18 | 106 | 23.33 | 231 | 50.84 | 206 | 45.34 |
| **Uberlândia** | 961202 | 42 | 4.37 | 456 | 47.44 | 1404 | 146.07 | 2902 | 301.91 |

Source: SIH/SUS (2021–2024); IBGE Population Estimates (2024).
Rate = (number of hospitalizations ÷ population of the Immediate Geographic Region) × 100000

Despite regional variability, most IGRs exhibited a progressive increase in hospitalizations over the study period, with the most pronounced growth occurring between 2023 and 2024. Santa Cruz do Sul displayed a non-linear pattern, with an early peak in 2021, a subsequent decline, and a renewed increase in 2024. Other regions, such as Passo Fundo and Santa Maria, began with very low hospitalization rates in 2021–2022 but rose to moderate levels by 2023–2024, reflecting a delayed but evident escalation.

As shown in **Table 3**, most IGRs displayed higher dengue-related search activity in the later years of the study period compared with 2021. Although the trajectory was not strictly linear, many regions showed a general upward tendency over time, with notable increases in 2022 and again in 2024. This temporal pattern is consistent with the national hospitalization profile, which also registered higher counts in these two years.

**Table 3.** Dengue-related search activity by Immediate Geographic Region (2021–2024)

| IGRs | Physicians<br>2021–<br>2024* | 2021 |  | 2022 |  | 2023 |  | 2024 |  |
| --- | --- | --- | --- | --- | --- | --- | --- | --- | --- |
|  |  | N | Rate | N | Rate | N | Rate | N | Rate |
| **Alegre** | 1066 | 10 | 0.94 | 44 | 4.13 | 56 | 5.25 | 96 | 9.01 |
| **Belo Horizonte** | 76259 | 2296 | 3.01 | 5502 | 7.21 | 9302 | 12.20 | 11802 | 15.48 |
| **Campina Grande** | 6924 | 176 | 2.54 | 277 | 4.00 | 102 | 1.47 | 289 | 4.17 |
| **Campos dos Goytacazes** | 2704 | 22 | 0.81 | 56 | 2.07 | 90 | 3.33 | 119 | 4.40 |
| **Catalão** | 922 | 14 | 1.52 | 69 | 7.48 | 37 | 4.01 | 139 | 15.08 |
| **Cruz Alta** | 1026 | 12 | 1.17 | 65 | 6.34 | 97 | 9.45 | 149 | 14.52 |
| **Distrito Federal** | 31532 | 2586 | 8.20 | 4927 | 15.63 | 3153 | 10.00 | 4808 | 15.25 |
| **Frederico Westphalen** | 779 | 15 | 1.93 | 89 | 11.42 | 47 | 6.03 | 199 | 25.55 |
| **Ijuí** | 1630 | 27 | 1.66 | 147 | 9.02 | 206 | 12.64 | 297 | 18.22 |
| **Juiz de Fora** | 8180 | 91 | 1.11 | 147 | 1.80 | 309 | 3.78 | 711 | 8.69 |
| **Linhares** | 2513 | 47 | 1.87 | 53 | 2.11 | 147 | 5.85 | 197 | 7.84 |
| **Maringá** | 8984 | 172 | 1.91 | 664 | 7.39 | 501 | 5.58 | 753 | 8.38 |
| **Marília** | 5296 | 65 | 1.23 | 186 | 3.51 | 188 | 3.55 | 325 | 6.14 |
| **Oliveira** | 1806 | 18 | 1.00 | 71 | 3.93 | 140 | 7.75 | 129 | 7.14 |
| **Passo Fundo** | 3274 | 30 | 0.92 | 158 | 4.83 | 229 | 6.99 | 283 | 8.64 |
| **Passos** | 1895 | 10 | 0.53 | 116 | 6.12 | 171 | 9.02 | 198 | 10.45 |
| **Pirapora** | 591 | 6 | 1.02 | 33 | 5.58 | 228 | 38.58 | 81 | 13.71 |
| **Porto Alegre** | 45502 | 930 | 2.04 | 5181 | 11.39 | 3171 | 6.97 | 5421 | 11.91 |
| **Ribeirão Preto** | 16605 | 189 | 1.14 | 442 | 2.66 | 500 | 3.01 | 1028 | 6.19 |
| **Rio de Janeiro** | 98975 | 2653 | 2.68 | 5161 | 5.21 | 8161 | 8.25 | 11130 | 11.25 |
| **Salvador** | 30866 | 1335 | 4.33 | 2635 | 8.54 | 2860 | 9.27 | 3882 | 12.58 |
| **Santa Cruz do Sul** | 2880 | 83 | 2.88 | 184 | 6.39 | 107 | 3.72 | 276 | 9.58 |
| **Santa Maria** | 3253 | 61 | 1.88 | 158 | 4.86 | 282 | 8.67 | 580 | 17.83 |
| **São Miguel do Oeste** | 939 | 11 | 1.17 | 92 | 9.80 | 71 | 7.56 | 137 | 14.59 |
| **São Paulo** | 182769 | 8556 | 4.68 | 11669 | 6.38 | 11012 | 6.03 | 28384 | 15.53 |
| **Uberaba** | 4419 | 64 | 1.45 | 260 | 5.88 | 310 | 7.02 | 355 | 8.03 |
| **Uberlândia** | 9906 | 146 | 1.47 | 620 | 6.26 | 965 | 9.74 | 1075 | 10.85 |

Source: Afya Whitebook Access and Engagement Data. Internal company platform. \* Physicians with at least one access to the Afya Whitebook® platform between 2021 and 2024. $N$ = number of dengue-related searches. Rate = $(N \div \text{physicians with at least one access to the Afya Whitebook®}) \times 100$

### 3.3 Correlation Analysis Between Hospitalizations, Climate Variables, and Digital Clinical Search Activity

The resulting correlation matrix revealed a clear contrast in the strength and consistency of associations across variable categories, as illustrated in **Figure 2**. Afya Whitebook® access demonstrated the strongest and most frequent positive correlation with dengue hospitalizations across IGRs.

**Figure 2.**
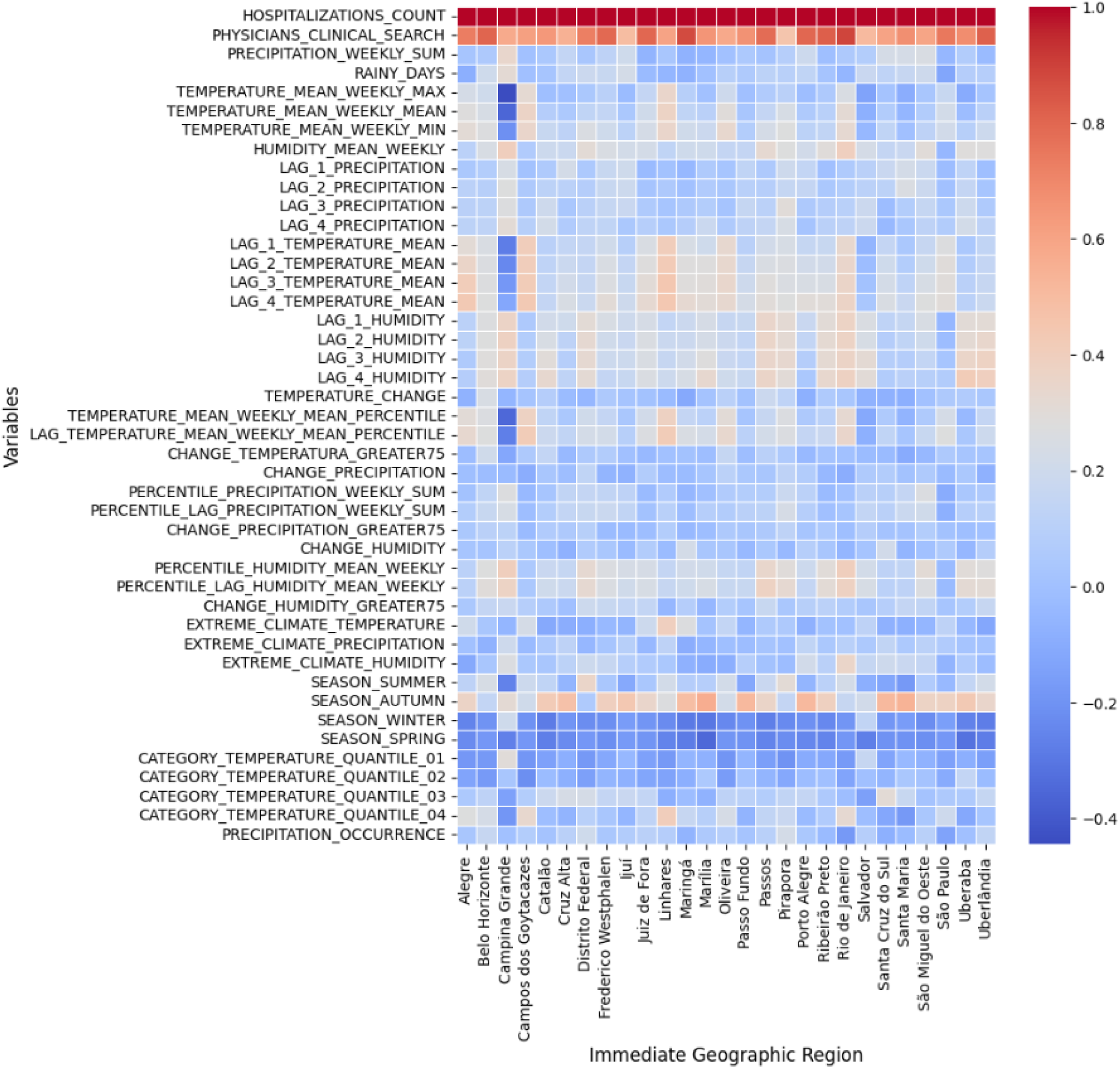
Correlation matrix showing pairwise correlations between dengue hospitalizations, Afya Whitebook® search activity, and climate-related predictors across the 27 Immediate Geographic Regions. Afya Whitebook® access demonstrates the strongest association with hospitalizations.

In contrast, climatic predictors displayed predominantly weak and heterogeneous correlations with dengue hospitalizations across IGRs. Humidity and temperature indicators showed low to moderate associations, with no consistent pattern across regions. Precipitation-related variables exhibited minimal and highly variable correlations, without any clear or recurrent signal in most regions.

Time-series curves of Afya Whitebook® searches and dengue hospitalizations further supported these findings, as illustrated in **Figure 3**. In nearly all IGRs, peaks in dengue-related search activity preceded or coincided with increases in hospitalizations.

**Figure 3.**
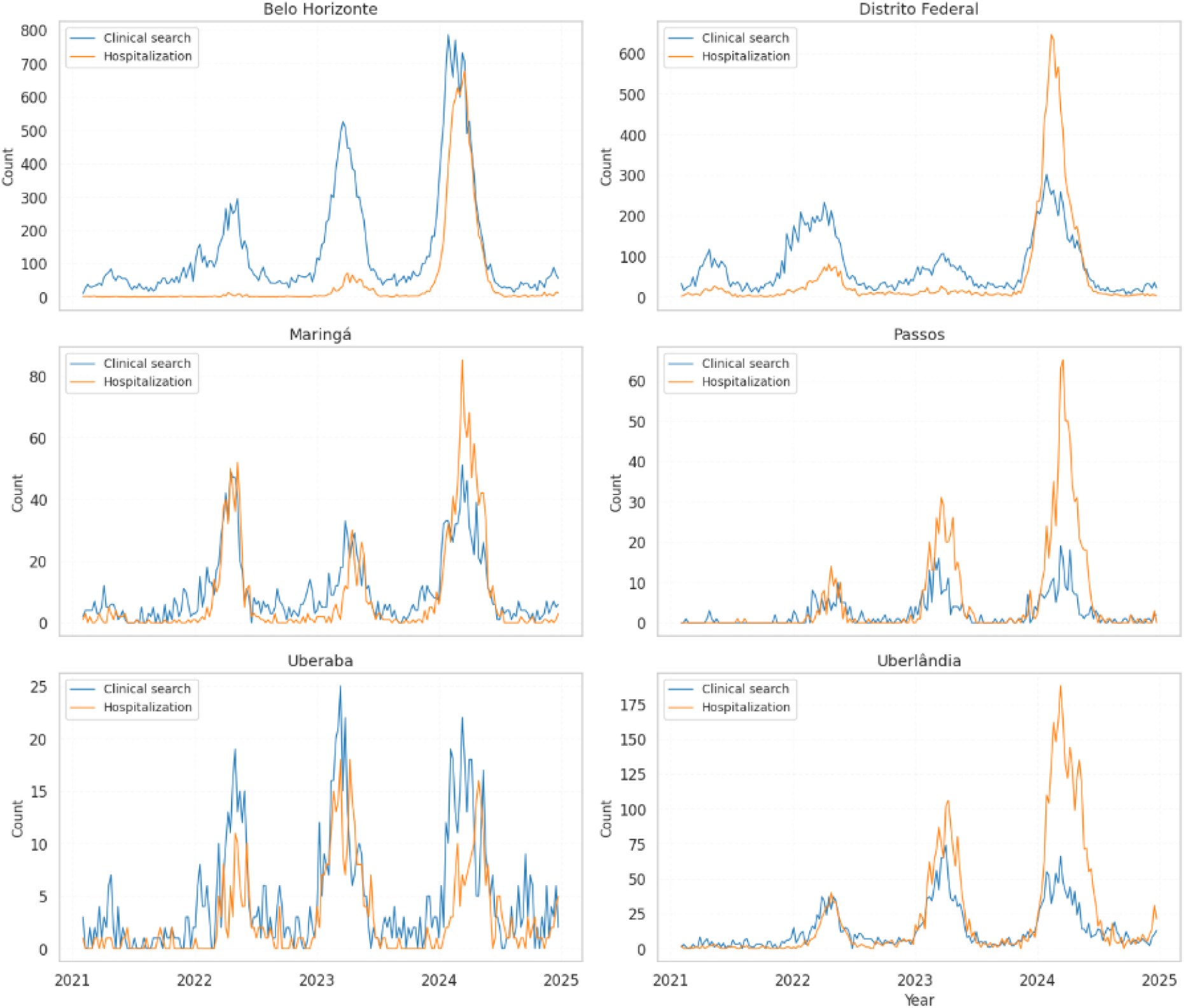
Weekly dengue hospitalizations (orange line) and Afya Whitebook® dengue-related search activity (blue line) across Immediate Geographic Regions from 2021 to 2024. In this figure, six of the 27 IGRs were selected as representative examples for visualization. Peaks in clinical search activity typically precede or coincide with hospitalization increase, indicating early clinical engagement with dengue and supporting its use as a real-time digital indicator.

### 3.4 Selected Variables Across Immediate Geographic Regions (IGRs)

During the feature-selection procedure, we ensured that each IGR retained at least one predictor from the four predefined climatic categories—humidity, temperature, precipitation, and seasonality—reflecting their established relevance in dengue transmission dynamics. Despite this harmonized structure, the LSTM model selected markedly different climatic signatures across regions. Some IGRs were primarily influenced by short-term humidity fluctuations or temperature lags, whereas others showed stronger contributions from precipitation extremes, or accumulated rainfall.

The full set of predictors selected for each IGR, grouped by climatic category, is provided in **Supplementary Table S2**.

### 3.5 Predictive Performance of Climate, Digital, and Integrated Models

Under the Ideal-data model (Figure 4), the Climate model consistently achieved the lowest prediction errors across nearly all IGRs, displaying the smallest RMSE medians and the narrowest interquartile ranges. The Integrated model performed as the second-best approach, although its accuracy gains over climate alone were modest. In contrast, the Clinical-search model showed higher error levels and greater variability. Detailed RMSE values (mean ± standard error) for all IGRs, as well as the corresponding pairwise statistical comparisons between model configurations, are reported in **Supplementary Tables S3 and S5**.

**Figure 4.**
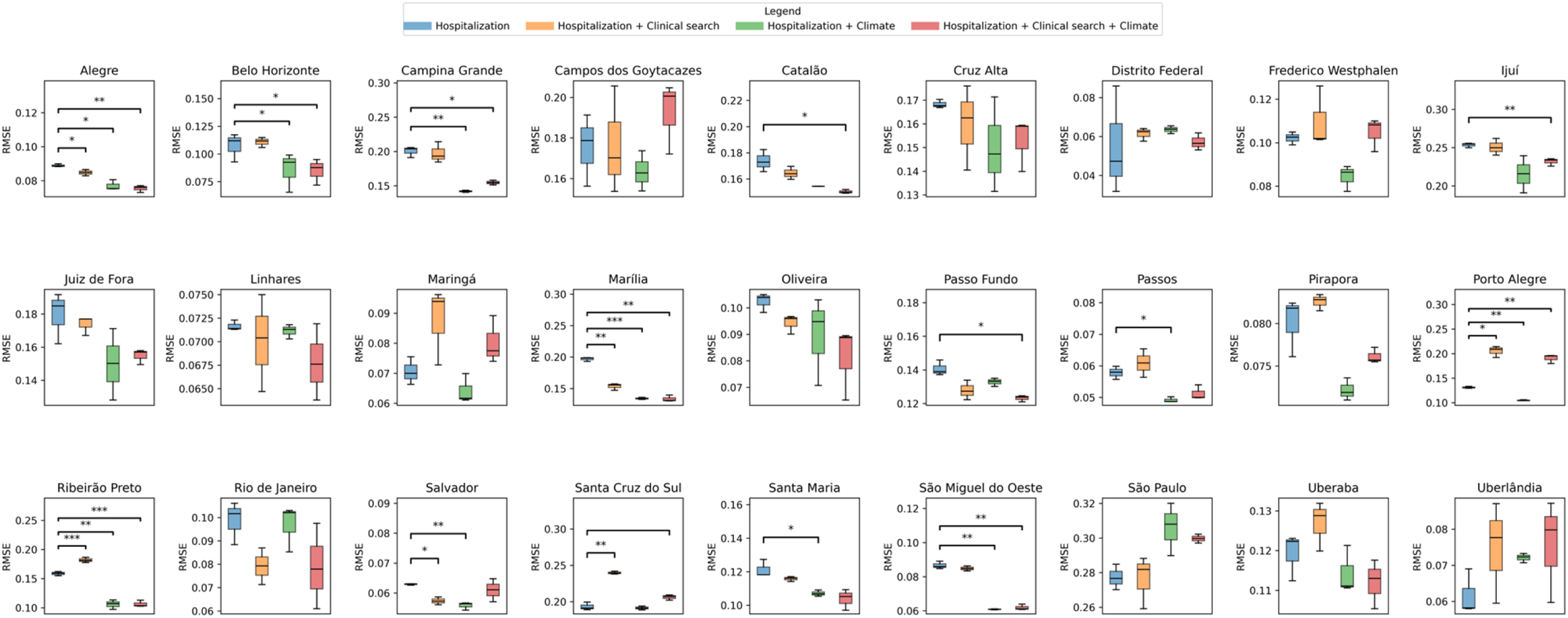
Distribution of RMSE values across the 27 Immediate Geographic Regions (IGRs) for the four modeling scenarios without hospitalization data delay: (i) LSTM- Hospitalization only, (ii) Clinical Search model (LSTM-Hospitalization + Clinical Search), (iii) Climate model (LSTM-Hospitalization + Climate), and (iv) Integrated model (LSTM- Hospitalization + Clinical Search + Climate). Each box represents the dispersion of RMSE scores across triplicate model runs for each IGR, allowing comparison of model accuracy under different predictor sets.

Under the Real-data model (Figure 5), where hospitalization data include reporting delays, the Integrated model consistently achieved the lowest prediction errors across most IGRs. Both the Climate and Clinical-search models displayed higher RMSE values, although the Clinical-search model showed relatively better performance than in the Ideal-data scenario. Climatic predictors remained informative but showed reduced accuracy relative to the no-delay condition. Consistent with this increased heterogeneity, a greater number of regions exhibited statistically significant pairwise differences between models in the Real-data scenario, as detailed in **Supplementary Tables S4 and S6**.

**Figure 5.**
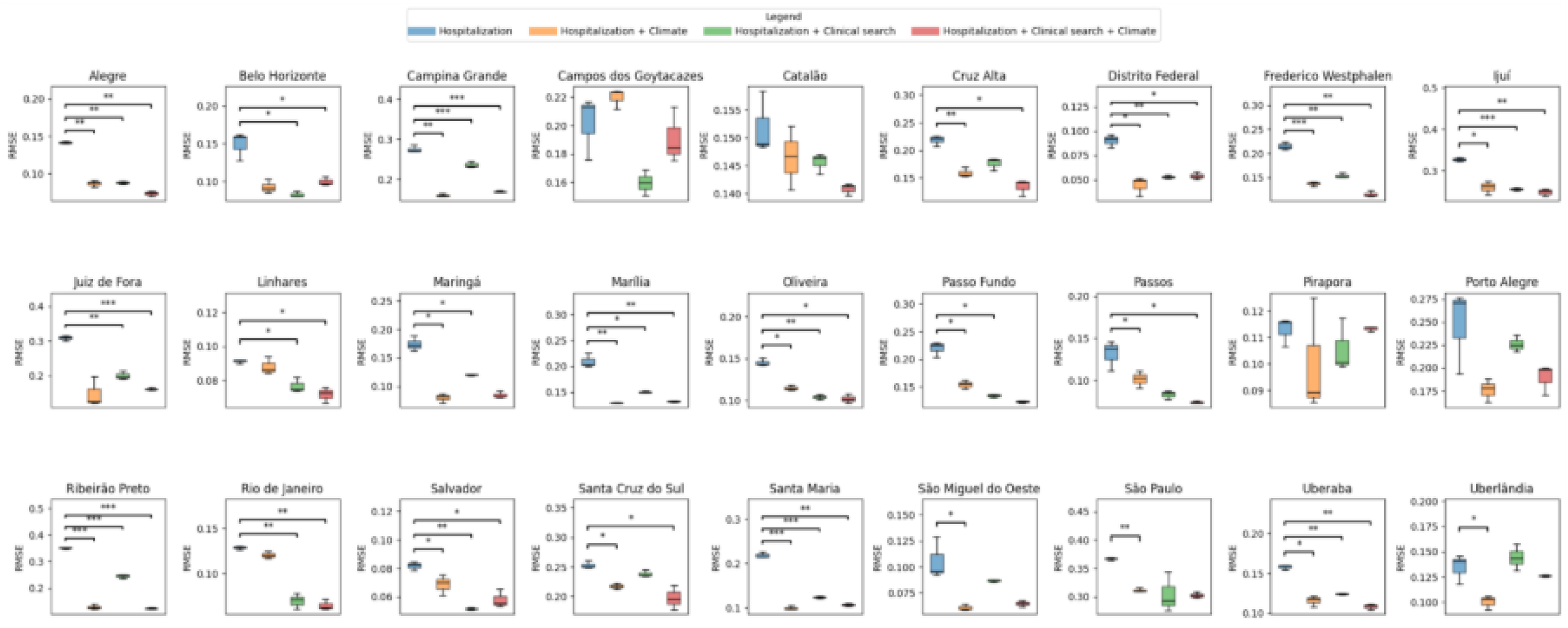
Distribution of RMSE values across the 27 Immediate Geographic Regions (IGRs) for the four modeling scenarios when incorporating realistic hospitalization data delays. The scenarios mirror those used in the no-delay analysis, enabling comparison of prediction performance under real-world reporting constraints

### 3.6 SHAP

When comparing SHAP patterns across scenarios (Figure 6; Supplementary Figures 7 and 8), a consistent shift emerged: the predictive influence of hospitalization counts decreased markedly in the Real-world model, reflecting the loss of temporal alignment introduced by reporting delays. As hospitalization became less informative, the relative importance of the clinical-search indicator increased, appearing more prominently across multiple IGRs. Although climatic predictors remained the most influential domain overall, their predominance varied substantially across regions—some IGRs were driven by humidity-related changes, others by temperature categories or precipitation lags—highlighting strong geographic heterogeneity.

**Figure 6.**
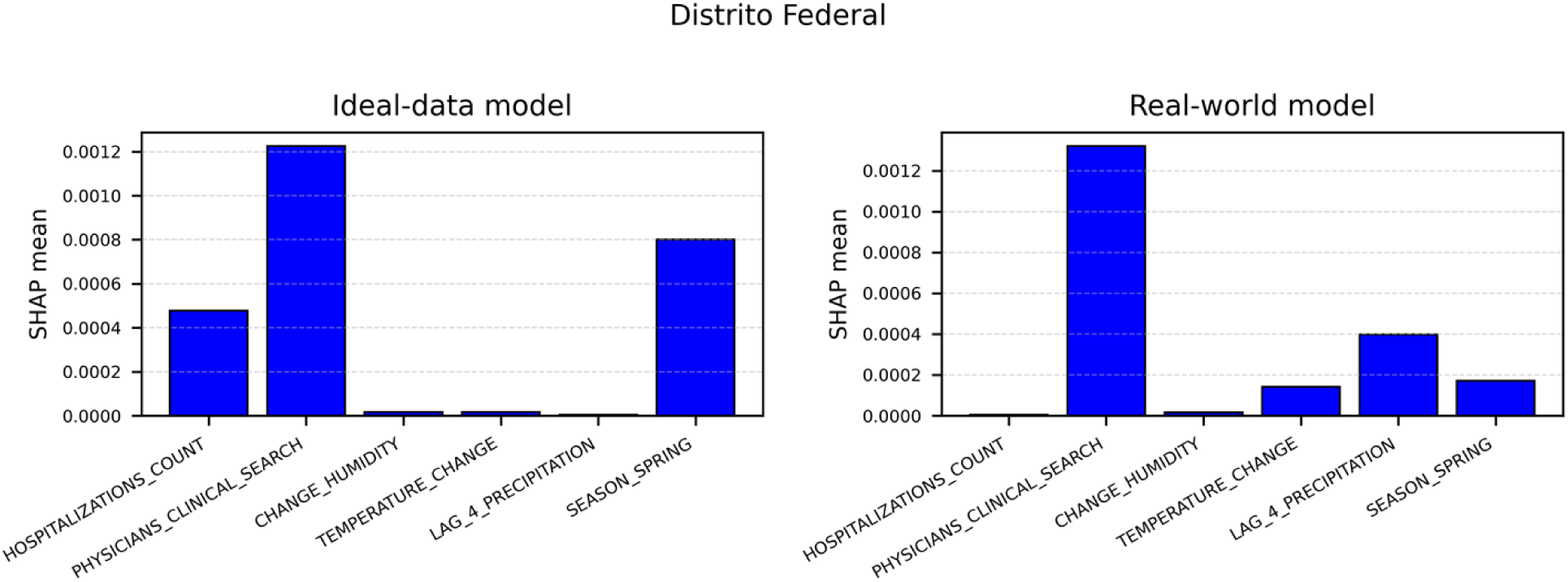
Across both models, the contribution of hospitalization counts dropped sharply under reporting delays. In contrast, the relative importance of the clinical-search indicator increased slightly in the Real-world scenario, reflecting its greater utility when hospitalization data becomes less informative.

## 4. Discussion

This study yielded three main findings. First, dengue-related searches on the Afya Whitebook® platform showed strong temporal alignment with hospitalizations, with search peaks often preceding or coinciding with increases in admissions. Second, climatic predictors varied substantially across IGRs, reflecting regional heterogeneity in environmental drivers. Third, under realistic reporting delays, the integrated model combining climate variables and digital search activity outperformed all other approaches, indicating that physician search behavior provides timely epidemiological information that compensates for the loss of predictive signal caused by delayed hospitalization data.

Between 2021 and 2024, clinical searches and hospitalizations increased across regions, with strong temporal alignment between the two. This indicates that physicians rapidly adjust their information-seeking behavior in response to changes in clinical demand, consistent with evidence that digital engagement can serve as an early indicator of infectious disease activity (13,14). Prior studies have shown that public digital indicators—such as Twitter activity—can forecast dengue trends by several weeks (25). By using physician search data rather than public-facing digital behavior, our study likely provides a more precise and clinically relevant digital signal, directly connected to medical decision-making and more suitable for operational forecasting.

Climatic variables contributed mainly through lagged effects, aligning with findings from Chen & Moraga (2025) that forecasting accuracy improves when models explicitly incorporate temporal delays in environmental exposures (11) In our study, contemporaneous associations between climate and hospitalizations were modest, but the LSTM consistently selected lagged temperature, humidity, and precipitation indicators as relevant inputs, and SHAP analyses confirmed their predictive influence. These results reinforce that climate is a key determinant of dengue dynamics, but its effects become detectable only when temporal structure and non-linear relationships are modeled explicitly.

Model comparisons further supported these patterns. In the ideal scenario without reporting delays, climate models achieved the lowest errors, consistent with the recognized environmental dependence of dengue transmission (4,5). When reporting delays were incorporated, predictive performance shifted: the integrated model became superior in most regions, and digital search data gained importance by providing real-time information that compensated for the diminished value of delayed hospitalization counts.

SHAP analyses clarified the interplay between predictors across scenarios. In the absence of reporting delays, climatic variables and recent hospitalization history dominated model contributions, whereas under delayed-reporting conditions, the influence of hospitalization history decreased sharply and digital search activity rose in relative importance. Together, these findings demonstrate that medium-range climatic signals and real-time behavioral indicators provide complementary information, enhancing the accuracy and operational value of forecasting models. This has practical implications for environmental and health surveillance: in settings where reporting delays are structural, incorporating digital search behavior into forecasting frameworks can improve situational awareness, support earlier public health responses, and strengthen regional early-warning systems. Similar benefits have been observed in other hybrid environmental–digital approaches used for dengue nowcasting in Brazil (26), reinforcing the potential of integrating clinical digital signals into surveillance tools.

Different limitations should be acknowledged when interpreting the generalizability of our model. The ecological design restricts inference to population-level associations and precludes individual causal interpretation. SIH/SUS hospitalization data may contain underreporting and reporting delays, potentially distorting temporal dengue patterns. Uneven INMET meteorological station coverage led to exclusions of regions with substantial data gaps, limiting nationwide applicability. Afya Whitebook® search patterns, while high-volume, represent only a subset of clinicians and may be biased toward younger, urban, or digitally engaged users. The absence of entomological or virological indicators (e.g., vector density or circulating serotypes) may also constrain the model’s sensitivity to short-term transmission shifts. Finally, LSTM models require large training datasets and remain partially opaque despite SHAP-based interpretability.

To mitigate these potential sources of bias, we implemented several measures. First, we restricted Afya Whitebook® data to verified physician accounts, reducing the risk of non-clinical or misclassified users and ensuring that digital indicators reflected genuine clinical activity. Second, we applied rigorous data-cleaning steps to the SIH/SUS time series and interpreted results only at the IGR level, where aggregation minimizes random fluctuations and reporting noise. Finally, we limited our analysis to IGRs with adequate climatic coverage, improving the reliability of environmental predictors.

This study demonstrates that clinician search behavior within a point-of care medical decision tool platform can serve as a timely and reliable digital indicator of dengue activity in Brazil. By integrating climatic conditions with real-time physician search data into our models, we consistently improve the forecasting of dengue hospitalizations, particularly when accounting for reporting delays. This integrated approach provides a more rapid and clinically grounded signal to support outbreak detection and public health decision-making. The next steps are to assess this framework’s generalizability across diverse settings and then evaluate its suitability for operational early-warning use, paving the way for its real-world application.

## 5. Supporting information

S1 Table. Climatic and clinical variables used in the forecasting models.

Description of all climatic, seasonal, and physician clinical-search variables, including units, derivation, and lagged indicators.

S2 Table. Predictors selected across Immediate Geographic Regions.

Climatic and digital predictors selected by the machine learning model across all Immediate Geographic Regions.

S1 Fig. Correlation between dengue hospitalizations, climate variables, and physician search activity.

Pairwise correlation analysis across regions.

S2 Fig. Time-series patterns of dengue hospitalizations and physician search activity.

Weekly trends showing the temporal alignment between clinical searches and hospitalizations.

S3 Table. Model performance under ideal-data conditions.

Root mean squared error values across regions for all predictive model configurations.

S4 Table. Statistical comparison of models under ideal-data conditions. Pairwise statistical tests comparing predictive performance.

S5 Table. Model performance under real-world reporting conditions.

Root mean squared error values across regions accounting for reporting delays.

S6 Table. Statistical comparison of models under real-world reporting conditions. Pairwise statistical tests comparing predictive performance under reporting delays.

S7 Fig. Feature importance under ideal-data conditions.

Relative contribution of predictors across regions based on model interpretation analyses.

S8 Fig. Feature importance under real-world reporting conditions. Changes in predictor importance when reporting delays are incorporated.

## Acknowledgements

We thank Beatriz Landiosi Teixeira for creating the graphical abstract and supporting the visual development of the study’s illustrative materials.

## Data availability

Hospitalization data from the Brazilian Hospital Information System (SIH/SUS) and meteorological data from the National Institute of Meteorology (INMET) are publicly available through the official portals listed in the references. Clinical-search data from the Afya Whitebook® platform contain proprietary and sensitive usage information and cannot be publicly released. These data can be made available upon reasonable request to the corresponding author, subject to approval by Afya®.

## Code availability

All code used for data processing, model development, and analysis is available in the project’s Jupyter notebooks. These materials can be provided upon reasonable request to the corresponding author.

